# After Reaching Its Highest Levels since 1950s, Incidence of Syphilis Among US Adults Declined in 2023

**DOI:** 10.1101/2024.04.02.24305225

**Authors:** Duy Do, Patricia J. Rodriguez, Samuel Gratzl, Brianna M. Goodwin Cartwright, Charlotte Baker, Nicholas L Stucky

**Affiliations:** Truveta, Incorporated, Bellevue, WA

## Abstract

Recent reports showed that the incidence of syphilis in the US reached a 70-year high. Using 2019-2023 data from Truveta, this study demonstrated that while the incidence of syphilis increased from 2020 to 2022, it started to decline in 2023. The decline was driven by population subgroups that are commonly considered higher-risk for syphilis infection. Findings also highlighted growing trends among lower-risk population subgroups.

## Introduction

Syphilis is a preventable and curable sexually transmitted infection (STI) caused by the bacteria Treponema pallidum [1]. Syphilis progresses through different stages, including primary, secondary, latent, and tertiary. Left untreated, syphilis is associated with life-threatening complications – including brain damage, deafness, blindness, fetal death, and an increased risk of acquiring other STIs [2]. These adverse health outcomes translate to substantial financial burdens for the US healthcare system, with an estimated total lifetime direct medical cost of $247 million in 2022 [3,4].

A recent report from the Centers for Disease Control and Prevention (CDC) suggested that the prevalence of syphilis in the US in 2022 reached its highest level since the 1950s – raising public concerns about the transmission of the disease and the health consequences of untreated syphilis, especially during the current shortage of penicillin [4,5]. The prevalence of primary and secondary (PS) syphilis – the most infectious stages of the disease – increased from a record low of 2.1 per 100,000 population in 2000 to 17.7 in 2022 [4,6]. Furthermore, the rate of PS syphilis increased for virtually all demographic subpopulations, with evidence of widening disparities in disease prevalence [4]. Between 2018 and 2022, the prevalence of PS syphilis increased by 335% for non-Hispanic American Indian or Alaska Native (AI/AN) persons, followed by 88.8% for Native Hawaiian or Pacific Islander (NH/PI) persons, compared to 70% for non-Hispanic white persons, highlighting broadening inequality in underlying risk factors for syphilis infection [4,7]. The rate of PS syphilis among women increased considerably (190%) between 2018 and 2022, compared to 44.1% for men [4], which may have implications for congenital syphilis and adverse pregnancy outcomes [8]. Given the resurgence of syphilis, the US Preventive Services Task Force recently reaffirmed its A-graded recommendation for syphilis screening in asymptomatic nonpregnant adolescents and adults [9].

Efforts aiming to contain the transmission of syphilis require the most recent data on disease incidence. While the CDC maintains a robust active surveillance program, their latest data on syphilis is from 2022. Understanding more-recent trends in syphilis infection may inform the design and implementation of effective public health programs. This study assessed monthly trends in the incidence of syphilis among US adults from January 2019 to December 2023, overall and stratified by demographic characteristics and risk factors.

## Methods

Syphilis incidence between 2019-2023 was estimated from a subset of Truveta Data – a continuously updated and linked database of electronic health records (EHR) from a collective of US healthcare systems. Data included information on patient demographics, encounters, diagnoses, medication, and diagnostic tests and results. Data was analyzed in March 2024. This study was exempt from Institutional Review Board approval because it used de-identified data.

The monthly incidence of syphilis was calculated from 2019 to 2023. In each month, the at-risk population included persons aged 18 and older with any medical encounter. Individuals were excluded if they had no medical encounters within one year before a given month or if they had any previously confirmed syphilis diagnosis. To ascertain incident syphilis (at any stage), individuals were required to have a diagnosis for syphilis during a medical encounter and reactive/positive results for a treponemal test and a non-treponemal test in the window of 30 days before to 14 days following their diagnosis. Individuals’ demographic characteristics included age (categorized as 18-34, 35-49, 50-64, and 65+), sex (female, male, unknown), race (Asian, Black, white, and others [AI/AN, NH/PI, and multiple races], unknown), Hispanic ethnicity, and census region of residence (Midwest, Northeast, South, West, unknown). Included syphilis risk factors were whether a person engaged in high-risk sexual activities, used Pre-Exposure Prophylaxis (PrEP), or had a previous HIV/AIDS diagnosis (see Table 1 for definitions and Table 2 for missing rates of these variables).

**Table 1:** Definitions for variables in the study.

|  |  |
| --- | --- |
| **Syphilis** | **ICD-10 codes:** A51*, A52*<br>**SNOMED CT codes:** 1107004, 3589003, 192008, 4082005, 4483005, 8555001, 10345003, 11338007, 12232008, 13095005, 13310005, 13731006, 16070004, 19206003, 20735004, 21523006, 22386003, 26039008, 26135000, 27460003, 28198007, 30080002, 31015008, 31137003, 36276008, 37430004, 37754005, 38523005, 39085002, 42770003, 44568006, 45058001, 46235002, 49923008, 50528008, 51928006, 51960003, 52414005, 54274001, 55768006, 58056005, 58227000, 59233003, 59307008, 59530001, 59934002, 60528006, 61612001, 62207008, 62861003, 63751007, 64102008, 66281009, 67125004, 67391006, 68764005, 69595007, 72083004, 75299005, 76272004, 77028001, 77939001, 80770009, 81339006, 82323002, 82355002, 82959004, 83883001, 85857008, 86028001, 87318008, 88943008, 91554004, 186833000, 186842007, 186846005, 186847001, 186850003, 186854007, 186861006, 186863009, 186867005, 186868000, 186875004, 186877007, 186878002, 186903006, 192647003, 193786000, 194907008, 194947001, 197305002, 197347003, 197348008, 197757004, 197966009, 198175009, 199154009, 199156006, 199157002, 199158007, 199159004, 202933002, 230152000, 230182006, 230563005, 230735006, 232313005, 232367004, 233849007, 234017002, 235032001, 235062007, 235064008, 235065009, 237446005, 237447001, 240552005, 240555007, 240556008, 240557004, 240558009, 240560006, 240562003, 240563008, 240564002, 240565001, 240566000, 240567009, 240568004, 240569007, 266125005, 266126006, 266127002, 266128007, 266130009, 266133006, 266136003, 278480000, 278481001, 302813001, 312934004, 312955002, 314840009, 315826004, 371237000, 402940004, 402941000, 402942007, 402943002, 402944008, 402945009, 402946005, 402947001, 402949003, 402950003, 402951004, 405635002, 410470003, 410478005, 444150000, 713251003, 735515000, 736686006, 19290004, 66887000, 86443005, 240554006 |
| **Treponemal lab test** | Microhemagglutination assay for T pallidum, T pallidum particle agglutination, T pallidum hemagglutination assay, Fluorescent treponemal antibody absorbed (FTA-ABS) test, Treponemal chemoluminescence immunoassays, Treponemal enzyme immunoassays<br>**LOINC codes:** 103199-6, 11597-2, 13288-6, 17723-8, 17724-6, 17725-3, 17726-1, 17727-9, 17728-7, 17729-5, 22585-4, 22586-2, 22587-0, 22588-8, 22589-6, 22590-4, 22591-2, 22592-0, 22593-8, 22594-6, 24110-9, 24312-1, 26009-1, 26658-5, 29310-0, 34147-9, 34382-2, 34954-8, 39015-3, 40679-3, 40680-1, 41122-3, 41163-7, 47051-8, 47063-3, 47236-5, 47237-3, 47238-1, 47511-1, 47512-9, 47514-5, 49799-0, 49800-6, 50689-9, 50695-6, 51474-5, 51475-2, 51838-1, 51839-9, 53605-2, 5392-6, 5393-4, 5394-2, 57032-5, 58031-6, 58751-9, 63464-2, 6561-5, 6562-3, 6580-7, 68502-4, 69946-2, 71793-4, 73752-8, 75175-0, 75190-9, 76766-5, 8041-6, 91846-6, 96611-9, 98212-4, 98213-2, 98214-0, 98215-7, 98216-5, 98217-3, 98218-1, 98219-9, 98220-7, 98221-5, 98222-3, 98223-1, 98224-9, 9826-9 |
| **Non-treponemal lab tests** | Rapid plasma reagin (RPR), Venereal Disease Research Laboratory (VDRL)<br>**LOINC codes:** 11084-1, 14904-7, 20507-0, 20508-8, 22459-2, 22460-0, 22461-8, 22462-6, 22464-2, 31146-4, 31147-2, 43813-5, 46203-6, 47235-7, 47476-7, 50690-7, 51783-9, 5289-4, 5290-2, 5291-0, 5292-8, 75195-8, 87925-4, 98212-4, 98214-0 |
| **High-risk sexual activities** | **ICD-10 codes:** Z72.51, Z72.52, Z72.53<br>**SNOMED CT codes:** 102947004, 316236005, 1220567008, 702479006 |
| **PrEP use**<br>A patient was defined as a PrEP user if in the 1 year prior, they met all following criteria:<br>(a) Filled any prescription for TDF/FTC (oral) or cabotegravir (injectable) for PrEP without a third antiviral being filled on the same day. Days-supply for TDF/FTC must be >28 days, otherwise we assume TDF/FTC was used for PEP.<br>(b) Had no diagnosis for HIV/AIDS and didn't fill any prescription for HIV/AIDS treatment (either a combined therapy of TDF/FTC and a third antiviral or cabotegravir and rilpivirine, or a one-dose therapy).<br>(c) Had no diagnosis for Hepatitis B. | **TDF/FTC (oral):** tenofovir disoproxil fumarate / emtricitabine.<br>**PrEP (injectable):** cabotegravir.<br>**Antiviral for anti-HIV/AIDS when used in combination with TDF/FTC:** lamivudine, efavirenz, zidovudine, lopinavir/ritonavir, raltegravir, dolutegravir, darunavir, ritonavir, abacavir, rilpivirine, etravirine, doravirine, nevirapine, tipranavir, atazanavir, and cobicistat.<br>**HIV diagnosis:**<br>**ICD-10 codes:** B20*, B97.35, O98.7*, Z21*<br>**SNOMED CT codes:** 10755671000119100, 111880001, 15928141000119100, 186706006, 186707002, 186708007, 230180003, 230598008, 235009000, 235726002, 236406007, 240103002, 276665006, 276666007, 281388009, 315019000, 397763006, 398329009, 402901009, 405631006, 40780007, 414376003, 414604009, 416729007, 420244003, 420281004, 420302007, 420308006, 420321004, 420384005, 420395004, 420403001, 420452002, 420524008, 420543008, 420544002, 420549007, 420554003, 420614009, 420658009, 420691000, 420718004, 420721002, 420764009, 420774007, 420787001, 420801006, 420818005, 420877009, 420900006, 420938005, 420945005, 421020000, 421023003, 421047005, 421077004, 421102007, 421230000, 421272004, 421283008, 421312009, 421315006, 421394009, 421403008, 421415007, 421431004, 421454008, 421460008, 421508002, 421529006, 421571007, 421597001, 421660003, 421671002, 421706001, 421708000, 421710003, 421766003, 421827003, 421851008, 421874007, 421883002, 421929001, 421983003, 421998001, 422003001, 422012004, 422089004, 422127002, 422136003, 422177004, 422189002, 422194002, 422282000, 422337001, 442134007, 442537007, 48794007, 52079000, 62479008, 697904001, 697965002, 700053002, 713260006, 713275003, 713278001, 713297001, 713298006, 713299003, 713300006, 713316008, 713318009, 713320007, 713325002, 713339002, 713340000, 713341001, 713342008, 713349004, 713444005, 713445006, 713446007, 713483007, 713484001, 713487008, 713488003, 713489006, 713490002, 713491003, 713497004, 713503007, 713504001, 713505000, 713506004, 713507008, 713508003, 713510001, 713511002, 713523008, 713526000, 713527009, 713530002, 713531003, 713532005, 713533000, 713543002, 713544008, 713545009, 713546005, 713570009, 713571008, 713572001, 713695001, 713696000, 713718006, 713722001, 713729005, 713730000, 713731001, 713732008, 713733003, 713734009, 713844000, 713845004, 713880000, 713881001, 713887002, 713897006, 713964006, 713967004, 714083007, 714464009, 719522009, 721166000, 722557007, 733834006, 733835007, 735521001, 735522008, 735523003, 735524009, 735525005, 735526006, 735527002, 735528007, 765434008, 771119002, 771126002, 771127006, 79019005, 80191000119101, 81000119104, 86406008, 87117006, 90681000119107, 90691000119105, 91947003, 91948008<br>**Hepatitis B diagnosis:**<br>**ICD-10 codes:** B16.0*, B16.1*, B16.2, B16.9*, B17.0*, B18.0*, B18.1*, B19.10, B19.11<br>**SNOMED CT codes:** 1116000, 111891008, 13265006, 153091000119109, 165806002, 186624004, 186626002, 186639003, 235864009, 235865005, 235869004, 235871004, 26206000, 271511000, 313234004, 38662009, 406117000, 424099008, 424340000, 442134007, 442374005, 446698005, 50167007, 53425008, 60498001, 61977001, 66071002, 76795007, 838380002 |
| **HIV/AIDS**<br>A patient was defined as having HIV/AIDS if they in the 1 year prior, they met any of the following criteria:<br>(a) Had any diagnosis for HIV/AIDS. | See the "PrEP use for prevention" section for definitions of TDF/FTC, antiviral therapies for anti-HIV/AIDS, and diagnosis for HIV/AIDS<br>One-dose HIV/AIDS treatment:<br>• Efavirenz / emtricitabine / tenofovir<br>• Bictegravir / emtricitabine / tenofovir alafenamide<br>• Emtricitabine / rilpivirine / tenofovir<br>• Doravirine / lamivudine / tenofovir disoproxil<br>• Dolutegravir / lamivudine |
| (b) Filled any prescription for TDF/FTC (oral) or cabotegravir (injectable) for PrEP with a third antiviral being filled on the same day. Days-supply for TDF/FTC must be >28 days, otherwise we assume TDF/FTC was used for PEP. | • Cobicistat / elvitegravir / emtricitabine / tenofovir<br>• Dolutegravir / rilpivirine<br>• Emtricitabine / rilpivirine / tenofovir<br>• Cobicistat / darunavir / emtricitabine / tenofovir alafenamide<br>• Efavirenz / lamivudine / tenofovir disoproxil<br>• Cobicistat / elvitegravir / emtricitabine / tenofovir<br>• Abacavir / dolutegravir / lamivudine |
| (c) Filled any prescription for one-dose HIV/AIDS treatment |  |

**Table 2:** Characteristics of US adults with incident syphilis (at any stage), 2019-2023.

|  |  | By calendar year of first-recorded syphilis infection |  |  |  |  |
| --- | --- | --- | --- | --- | --- | --- |
|  | Overall<br>N = 10,382 | 2019<br>N = 1,614 | 2020<br>N = 1,542 | 2021<br>N = 2,173 | 2022<br>N = 2,541 | 2023<br>N = 2,512 |
|  |  | n (column %) unless otherwise noted |  |  |  |  |
| Age (years),<br>mean (SD) | 41.1<br>(15.3) | 42.4<br>(16.1) | 41.1<br>(15.3) | 40.7<br>(15.3) | 40.5<br>(14.9) | 41.3<br>(15.2) |
| Age group |  |  |  |  |  |  |
| 18-34 | 4,901<br>(47.2%) | 720<br>(44.6%) | 734<br>(47.6%) | 1,056<br>(48.6%) | 1,208<br>(47.5%) | 1,183<br>(47.1%) |
| 35-49 | 2,848<br>(27.4%) | 437<br>(27.1%) | 410<br>(26.6%) | 583<br>(26.8%) | 717<br>(28.2%) | 701<br>(27.9%) |
| 50-64 | 1,866<br>(18.0%) | 313<br>(19.4%) | 284<br>(18.4%) | 385<br>(17.7%) | 450<br>(17.7%) | 434<br>(17.3%) |
| 65+ | 767<br>(7.4%) | 144<br>(8.9%) | 114<br>(7.4%) | 149<br>(6.9%) | 166<br>(6.5%) | 194<br>(7.7%) |
| Sex |  |  |  |  |  |  |
| Female | 3,823<br>(36.8%) | 428<br>(26.5%) | 505<br>(32.7%) | 818<br>(37.6%) | 1,018<br>(40.1%) | 1,054<br>(42.0%) |
| Male | 6,446<br>(62.1%) | 1,170<br>(72.5%) | 1,025<br>(66.5%) | 1,338<br>(61.6%) | 1,485<br>(58.4%) | 1,428<br>(56.8%) |
| Unknown | 113<br>(1.1%) | 16<br>(1.0%) | 12<br>(0.8%) | 17<br>(0.8%) | 38<br>(1.5%) | 30<br>(1.2%) |
| Race |  |  |  |  |  |  |
| Asian | 212<br>(2.0%) | 40<br>(2.5%) | 31<br>(2.0%) | 32<br>(1.5%) | 60<br>(2.4%) | 49<br>(2.0%) |
| Black | 4,044<br>(39.0%) | 740<br>(45.8%) | 607<br>(39.4%) | 862<br>(39.7%) | 954<br>(37.5%) | 881<br>(35.1%) |
| White | 4,279<br>(41.2%) | 578<br>(35.8%) | 658<br>(42.7%) | 895<br>(41.2%) | 1,082<br>(42.6%) | 1,066<br>(42.4%) |
| Other race<sup>b</sup> | 1,072<br>(10.3%) | 135<br>(8.4%) | 140<br>(9.1%) | 226<br>(10.4%) | 274<br>(10.8%) | 297<br>(11.8%) |
| Unknown | 775<br>(7.5%) | 121<br>(7.5%) | 106<br>(6.9%) | 158<br>(7.3%) | 171<br>(6.7%) | 219<br>(8.7%) |
| Ethnicity |  |  |  |  |  |  |
| Not Hispanic | 6,401<br>(61.7%) | 1,055<br>(65.4%) | 962<br>(62.4%) | 1,331<br>(61.3%) | 1,559<br>(61.4%) | 1,494<br>(59.5%) |
| Hispanic | 1,392<br>(13.4%) | 179<br>(11.1%) | 164<br>(10.6%) | 286<br>(13.2%) | 391<br>(15.4%) | 372<br>(14.8%) |
| Unknown | 2,589<br>(24.9%) | 380<br>(23.5%) | 416<br>(27.0%) | 556<br>(25.6%) | 591<br>(23.3%) | 646<br>(25.7%) |
| Census region |  |  |  |  |  |  |
| Midwest | 2,400<br>(23.1%) | 494<br>(30.6%) | 416<br>(27.0%) | 554<br>(25.5%) | 509<br>(20.0%) | 427<br>(17.0%) |
| Northeast | 592<br>(5.7%) | 101<br>(6.3%) | 98<br>(6.4%) | 111<br>(5.1%) | 147<br>(5.8%) | 135<br>(5.4%) |
| South | 2,952<br>(28.4%) | 396<br>(24.5%) | 375<br>(24.3%) | 572<br>(26.3%) | 800<br>(31.5%) | 809<br>(32.2%) |
| West | 2,291<br>(22.1%) | 290<br>(18.0%) | 294<br>(19.1%) | 470<br>(21.6%) | 596<br>(23.5%) | 641<br>(25.5%) |
| Unknown | 2,147<br>(20.7%) | 333<br>(20.6%) | 359<br>(23.3%) | 466<br>(21.4%) | 489<br>(19.2%) | 500<br>(19.9%) |
| **Used PrEP for prevention last 1 year** |  |  |  |  |  |  |
| No | 10,109<br>(97.4%) | 1,577<br>(97.7%) | 1,504<br>(97.5%) | 2,114<br>(97.3%) | 2,466<br>(97.0%) | 2,448<br>(97.5%) |
| Yes | 273<br>(2.6%) | 37<br>(2.3%) | 38<br>(2.5%) | 59<br>(2.7%) | 75<br>(3.0%) | 64<br>(2.5%) |
| **High-risk sexual activities last 1 year** |  |  |  |  |  |  |
| No | 9,742<br>(93.8%) | 1,502<br>(93.1%) | 1,440<br>(93.4%) | 2,027<br>(93.3%) | 2,396<br>(94.3%) | 2,377<br>(94.6%) |
| Yes | 640<br>(6.2%) | 112<br>(6.9%) | 102<br>(6.6%) | 146<br>(6.7%) | 145<br>(5.7%) | 135<br>(5.4%) |
| **HIV/AIDS** |  |  |  |  |  |  |
| No | 8,206<br>(79.0%) | 1,097<br>(68.0%) | 1,155<br>(74.9%) | 1,782<br>(82.0%) | 2,068<br>(81.4%) | 2,104<br>(83.8%) |
| Yes | 2,176<br>(21.0%) | 517<br>(32.0%) | 387<br>(25.1%) | 391<br>(18.0%) | 473<br>(18.6%) | 408<br>(16.2%) |

Note: Percentages may not sum up to 100% due to rounding.
a: Pearson's Chi-squared test.
b: American Indian or Alaska Native, Native Hawaiian or Pacific Islander, or multiple races.

The monthly incidence of syphilis was calculated by dividing the number of persons with incident syphilis by the number of at-risk persons. A Poisson model was used to estimate time trends in incidence of syphilis, with an offset variable for the monthly number of at-risk persons. All models included a fixed-effect variable indicating the first four months of the COVID-19 pandemic (March to June 2020) to account for reduced care-seeking behavior during this time. The independent variable of interest was a measure of continuous time, normalized to have a 0 to 1 range, using the following formula:

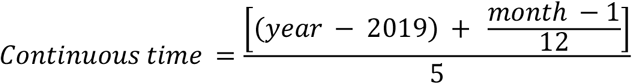

A one-unit change in the trend coefficient can be interpreted as a change in the incidence of syphilis across 5 years from the start (January 2019) to the end (December 2023) of the study period. The incidence rate ratio (IRR) was calculated by taking the exponential of the trend coefficient. Poisson model was then performed individually to test trend over the study period by population subgroups. This study followed the STROBE reporting guideline for cohort studies.

## Results

The study included 43,881,503 unique individuals spanning 422,384,538 person-months (mean age [SD]: 53.1 [18.8] years, 58.1% were women, 12.8% were Black, and 63.8% were white). From 2019 to 2023, 10,382 persons had incident syphilis (mean age [SD]: 41.1 [15.3] years, 36.8% were women, 39.0% were Black, and 41.2% were white). While a large proportion of patients with incident syphilis were male, Black, non-Hispanic, and living in the Midwest, their shares declined considerably over time. In contrast, the share of cases increased for those who were female, white, and those in the South or West regions (Table 2).

### Trends in Incidence of Syphilis Overall and by Demographic Characteristics

We observed an increasing trend in the overall incidence of syphilis among US adults over the 5-year period (IRR, 1.36; p-value < 0.001) (Figure 1a). Between January 2019 and May 2020, the incidence of syphilis declined from 2.4 to 1.4 per 100,000 persons, then rapidly grew until reaching its peak level of 3.2 in November 2022. It then dropped to 2.5 in December 2023.

**Figure 1:**
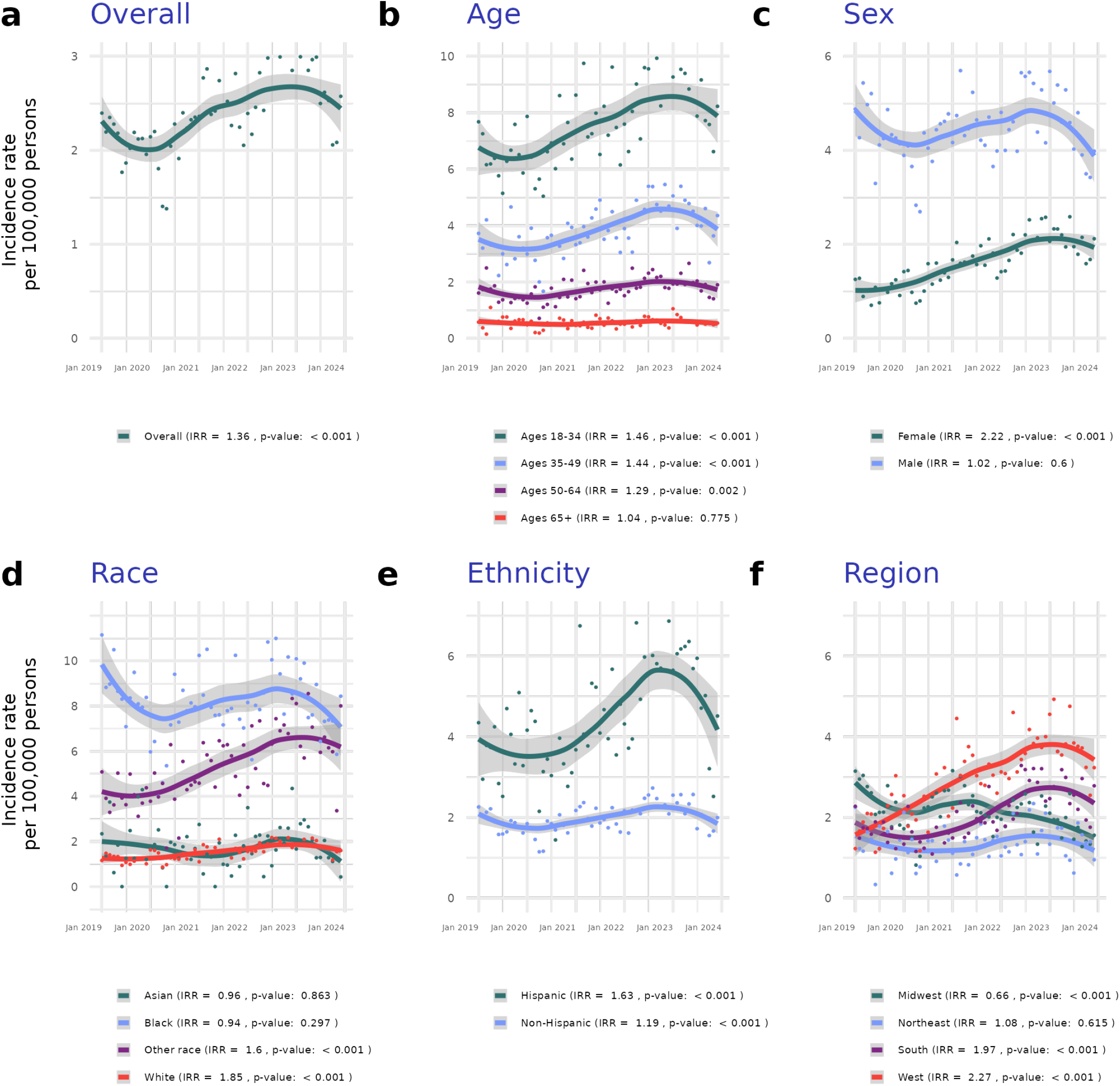
Trends in the incidence of syphilis (at any stage) among US adults ages 18 and older, overall and by selected demographic characteristics, 2019-2023. Note: IRR, incidence rate ratio. Gray areas depict 95% confidence interval around the fitted line. Trend in incidence of syphilis was smoothened using locally estimated scatterplot smoothing (LOESS), a non-parametric regression method that combines multiple linear regression models in a k-nearest-neighbor-based meta-model. A Poisson model was used to estimate time trends in the incidence of syphilis. The outcome variable was a monthly count of persons with incident syphilis. An offset variable for the monthly number of at-risk persons was included. A fixed-effect variable indicating the first four months of the COVID-19 pandemic (March to June 2020) was included to account for any effect of the pandemic and lockdown on care-seeking behavior. The independent variable of interest was a measure of continuous time, normalized to have a 0 to 1 range. A one-unit change in the trend coefficient is interpreted as a change in the incidence of syphilis across the 5-year period from the start (January 2019) to the end (December 2023) of the study period.

Trends in the incidence of syphilis varied by demographic characteristics (Figures 1b to 1f). The incidence increased from 2020 to 2022 for all age groups then declined in 2023, with younger adults experiencing a higher incidence rate during this period. From 2019 to 2023, trend in incident syphilis was relatively flat for men (IRR, 1.02; p-value = 0.60), while it increased more than two-folds for women (IRR, 2.22; p-value < 0.001). When stratifying by race, Black individuals had the highest rate of syphilis and white and Asian individuals had the lowest.

Nonetheless, trend in the incidence of syphilis was non-significant for Black individuals (IRR, 0.94; p-value = 0.30), while it increased for those identified as white (IRR, 1.85; p-value < 0.001) and other (IRR, 1.60; p-value < 0.001). Compared with non-Hispanic individuals (IRR, 1.19; p-value < 0.001), Hispanic adults had a higher level of and an increasing trend (IRR, 1.63; p-value < 0.001) in incident syphilis, although the incidence rate started to decline in 2023. Finally, trends in incident syphilis increased rapidly in 2019-2023 for those in the South (IRR, 1.97; p-value < 0.001) and West (IRR, 2.27; p-value < 0.001) regions, while it declined for those in the Midwest (IRR, 0.66; p-value < 0.001).

### Trends in Incidence of Syphilis by Risk Factors

Levels and trends in incident syphilis differed by risk factors (Figure 2a to 2c). Overall, trends in incident syphilis increased during the study period for those without evidence of high-risk sexual activities (IRR, 1.39; p-value < 0.001), PrEP use (IRR, 1.40; p-value < 0.001), or HIV/AIDS (IRR, 1.75; p-value < 0.001), despite having relatively low incident levels. In contrast, trends in incident syphilis were stagnant or declined significantly among those with high-risk sexual activities (IRR, 1.01; p-value = 0.96), PrEP use (IRR, 0.63; p-value = 0.03), or HIV/AIDS (IRR, 0.36; p-value < 0.001).

**Figure 2:**
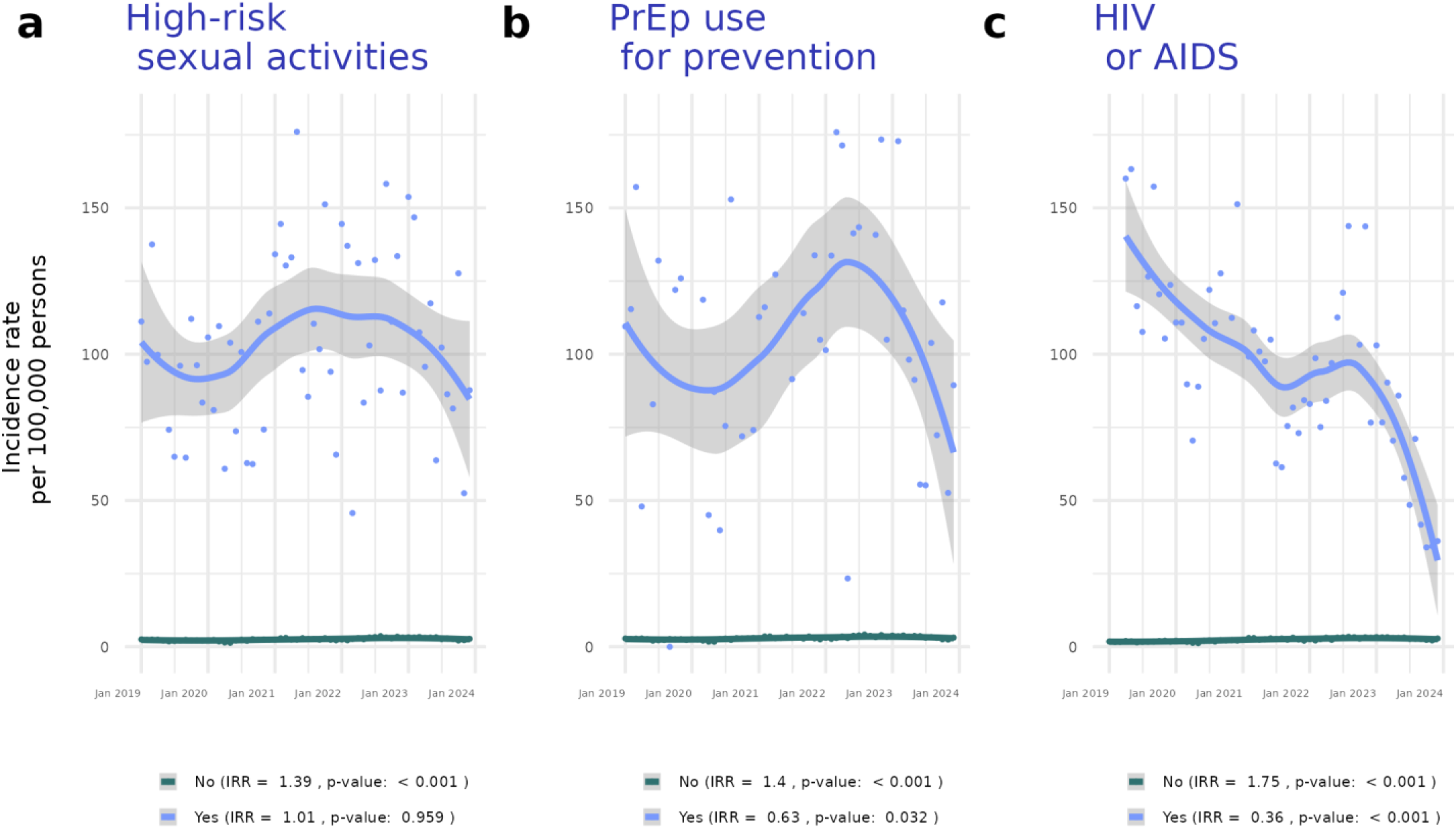
Trends in the incidence of syphilis (at any stage) among US adults ages 18 and older, by selected risk factors, 2019-2023. Note: IRR, incidence rate ratio. Gray areas depict 95% confidence interval around the fitted line. Trend in incidence of syphilis was smoothened using locally estimated scatterplot smoothing (LOESS), a non-parametric regression method that combines multiple linear regression models in a k-nearest-neighbor-based meta-model. A Poisson model was used to estimate time trends in the incidence of syphilis. The outcome variable was a monthly count of persons with incident syphilis. An offset variable for the monthly number of at-risk persons was included. A fixed-effect variable indicating the first four months of the COVID-19 pandemic (March to June 2020) was included to account for any effect of the pandemic and lockdown on care-seeking behavior. The independent variable of interest was a measure of continuous time, normalized to have a 0 to 1 range. A one-unit change in the trend coefficient is interpreted as a change in the incidence of syphilis across the 5-year period from the start (January 2019) to the end (December 2023) of the study period.

## Conclusion

Although the incidence of syphilis accelerated rapidly following the first few months of the COVID-19 pandemic, it reached an inflection point in November 2022, with a return to the pre-pandemic level by December 2023. The recent decline was primarily driven by individuals commonly considered high-risk for syphilis infection [6] – including men, younger adults, Black and Hispanic individuals, and those with high-risk sexual activities, PrEP use, or HIV/AIDS.

This study also highlighted growing trends in the incidence of syphilis among population subgroups that are generally considered low-risk [6] – namely women, individuals identified as white or ‘other’ (AI/AN, NH/PI, and multiple races), and those without evidence of high-risk sexual activities, PrEP use, or HIV/AIDS. Similar to other studies using EHR data [10], this study was subject to several limitations, such as the inability to capture events that occurred outside of Truveta constituent healthcare systems. Therefore, incidence rates in this study were lower than those reported by the CDC [4], although the trends were similar. Nevertheless, the advantages of this study included using more-recent data in 2023 to show a decline in incident syphilis rates and growing trends among lower-risk population subgroups. Future studies should investigate changes in the underlying social conditions and risk factors that contribute to growing trends in incident syphilis among these populations.

## Data Availability

All data used in this study are publicly available to Truveta subscribers and may be accessed at studio.truveta.com

https://www.studio.truveta.com

## Contribution

Conceptualization: All authors. Methodology: All authors. Data acquisition: All authors. Data analysis: All authors. Interpretation: All authors. Initial draft: DD. Revisions: All authors. Final document review and approval for publication: All authors.

## Funding

No external funding was obtained for this work.

### Ethics approval and consent to participate

Normalized electronic health record data are de-identified by expert determination under the HIPAA Privacy Rule before being made available to researchers. In accordance with 46.101 Protection of Human Subjects, our study did not require Institutional Review Board approval because it used only deidentified medical records. All necessary patient/participant consent has been obtained and the appropriate institutional forms have been archived, and that any patient/participant/sample identifiers included were not known to anyone (e.g., hospital staff, patients or participants themselves) outside the research group so cannot be used to identify individuals.

### Consent for publication

Not applicable.

### Availability of Data and Materials

The data and code used in this study is available to all Truveta subscribers and may be accessed at http://studio.truveta.com.

